# Antimicrobial treatment impacts resistance in off-target populations of a nosocomial bacterial pathogen: a case-control study

**DOI:** 10.1101/2020.01.28.20019323

**Authors:** Clare L. Kinnear, Elsa Hansen, Meghan Forstchen, Andrew F. Read, Robert J. Woods

## Abstract

The antimicrobial resistance crisis has persisted despite broad attempts at intervention. Detailed studies of the population dynamics that lead to resistance could identify additional intervention points. An important driver of resistance in the most concerning antibiotic resistant pathogens can be selection imposed on bacterial populations that are not the intended target of antimicrobial therapy. Here we focus on the important nosocomial pathogen *Enterococcus faecium* in a hospital system where resistance to daptomycin is evolving despite standard interventions. We hypothesized that the intravenous use of daptomycin generates off-target selection for resistance in transmissible gastrointestinal (carriage) populations of *E. faecium*. We performed a case-control study in which the daptomycin resistance of *E. faecium* isolated from rectal swabs from daptomycin-exposed patients was compared to a control group of patients exposed to linezolid, a drug with similar indications. In the daptomycin-exposed group, daptomycin resistance of *E. faecium* from the off-target population was on average 50% higher than resistance in the control group (n=428 independent *E. faecium* clones from 22 patients). There was also greater phenotypic diversity in daptomycin resistance within daptomycin-exposed patients. Multiple samples over time were available from a subset of patients, and these demonstrate wide variability in temporal dynamics, from long-term maintenance of resistance to rapid return to sensitivity after daptomycin treatment stopped. Our results demonstrate that off-target gastrointestinal populations rapidly respond to intravenous antibiotic exposure. Gastrointestinal populations are the source for faecal transmission and so can be the driver for hospital-wide population level increases in resistance. Focusing on the off-target evolutionary dynamics may offer novel avenues to slow the spread of antibiotic resistance.

## Introduction

Antimicrobial resistance emerges and spreads in response to antimicrobial treatment [1]. For the microbial population being intentionally targeted by drug treatment, selective pressure favoring resistance is an unavoidable consequence of suppressing population growth. However, antimicrobial exposure is not limited to the site of infection and so can exert selective pressure on so-called off-target or bystander populations [2–4]. This off-target selective pressure is particularly problematic for colonizing opportunistic pathogens, which include the multidrug-resistant pathogens of greatest concern [5]. Antimicrobial pressure imposed on these colonizing organisms has no therapeutic benefits, but can select for antibiotic resistant infections in the treated patient, or for the transmission of resistant isolates to other patients [6]. Selection for resistance in gastrointestinal carriage populations has been demonstrated in multiple opportunistic pathogens including Enterobacteriaceae [7–12], *Enterococcus* [13] and Bacteroides [14]. In many situations, selection on off-target populations may be a major contributor to population level resistance. For example, it has been estimated that as much as 90% of drug exposure experienced by *Klebsiella* is off-target [15]. The evolutionary dynamics in these off-target populations are poorly understood. This fundamental knowledge gap limits the ability to evaluate novel resistance management strategies in the off-target population such as choice of antimicrobial spectrum, use of antibiotic combinations, optimized routes of administration and addition of novel adjuvant therapies [6].

The evolution of daptomycin resistance among vancomycin-resistant *Enterococcus faecium* (VR *E. faecium*) is a relevant and tractable system to study off-target selection. VR *E. faecium* is an important cause of hospital acquired infections [16]. It spends the bulk of its life history asymptomatically colonizing the gastrointestinal tract of its host, but colonization is a key risk factor for clinical infections [17]. Intrinsic and acquired multi-drug resistance are common in VR *E. faecium*, leaving daptomycin as one of the few remaining treatment options. Daptomycin is delivered exclusively as an intravenous formulation, with 6% of the drug being excreted in the feces [18], allowing the potential for off-target selection. Daptomycin resistance has been shown to arise within patients during treatment [19]. Additionally, a number of studies have reported prior daptomycin treatment as a key risk factor for infection with daptomycin resistant VRE [20–22], which is consistent with off-target selection. Finally, transmission of VR *E. faecium* is common in hospital settings, meaning resistance that arises in one patient may pose a threat to others.

We hypothesize that most transmitted daptomycin resistance in VR *E. faecium* is due to off-target selection in the gastrointestinal tract of patients. This hypothesis predicts that daptomycin exposure is associated with daptomycin resistance in the off-target population, which has not been demonstrated previously. If this hypothesis is true, it raises additional questions: is there pre-existing variation within patients prior to exposure on which selection can act; is there more variability within some patients than others; does daptomycin exposure result in a single resistance phenotype dominating the gut; and finally, is variation in resistance maintained over time?

To address these questions, we focus on an institution where daptomycin resistance in *E. faecium* has been observed to evolve within patients and across the whole hospital population [19,23]. We investigate the impact of daptomycin exposure on daptomycin resistance in *E. faecium* colonizing the intestinal tract by utilizing rectal swabs available from a prospective surveillance program. The organisms colonizing the intestinal tract are not the target of treatment, thus resistance in this population represents unintended, off-target evolution. We perform a case-control study, in which patients exposed to daptomycin are compared to patients exposed to a drug with similar indications, linezolid. The available samples allow us to isolate and measure resistance in multiple independent *E. faecium* colonies per patient swab sample. We quantify the impact of drug exposure on the mean and the distribution of phenotypes in the colonizing population, which gives unique insight the into potential mechanism of competition and transmission in this pathogen. Finally, where samples are available, we explore changes in resistance over time within patients.

## Results

During the calendar year 2016, 6,726 patients had a rectal swab to screen for VRE colonization, of these, 618 patients were positive for VRE. Treatment with daptomycin within this pool of patients is low (23/618 patients, Fig 1), partially due to a change in antibiotic use in 2015 away from daptomycin [23]. Fourteen patients met the inclusion criteria for cases with at least three doses (three days of therapy) of daptomycin in the six months prior to the positive swab (daptomycin group). A further 15 patients met the criteria for the control group with no known prior daptomycin treatment and at least six doses (three days of therapy) of linezolid (Fig 1, see methods for further details). The first sample from each patient to meet these criteria is considered the index sample for that patient.

**Fig 1.**
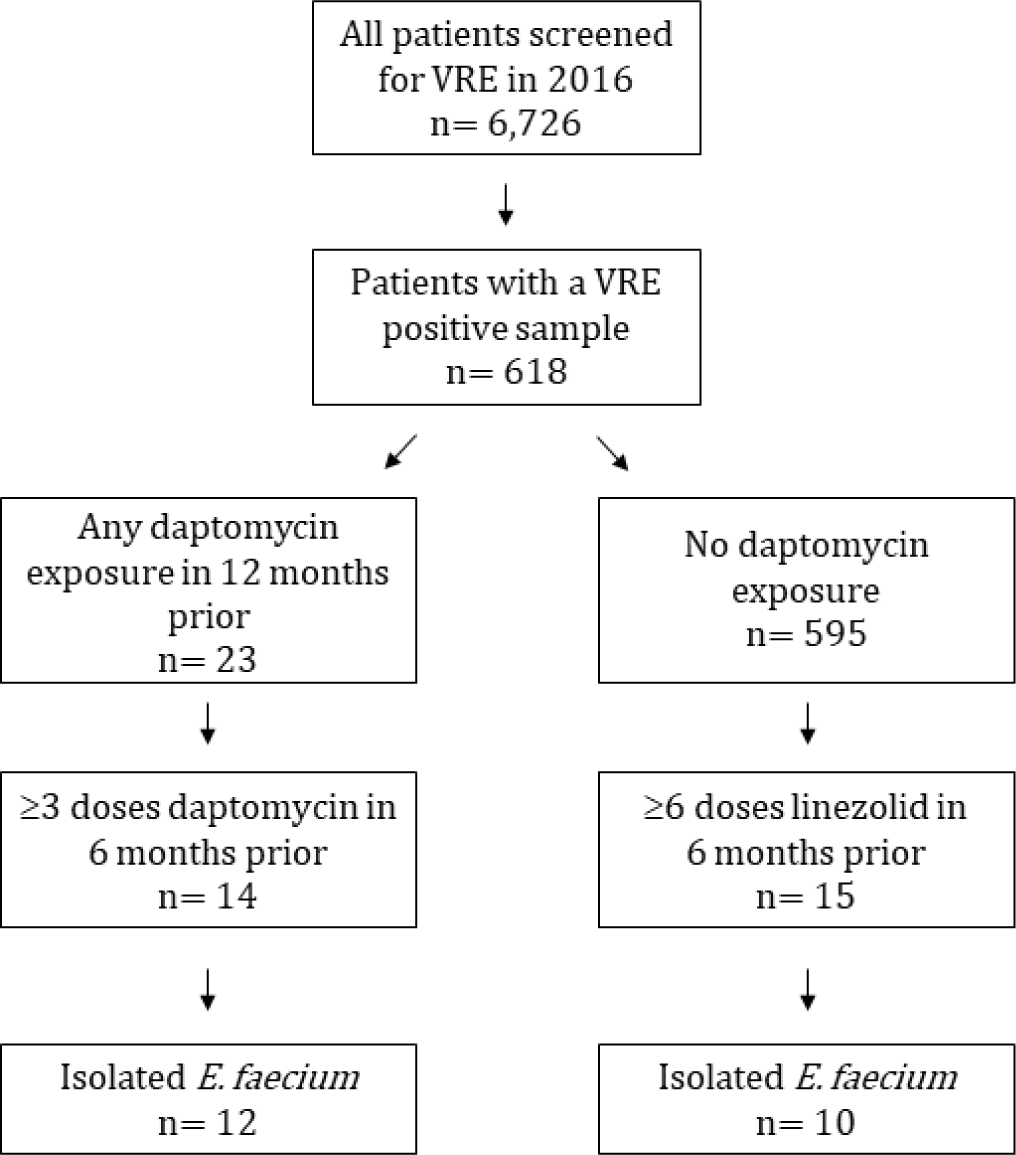
Identification of patients meeting the cases and control study definitions.

*Enterococcus* was isolated from all samples using Enterococcosel agar. While the inclusion criteria required a positive result for VR *E. faecium*, the collection protocol resulted in isolation of both vancomycin resistant and vancomycin susceptible *E. faecium*. For two of the daptomycin treated samples and five of the control samples, *E. faecium* was either absent or at a density (relative to other *Enterococcus* species) that did not allow isolation of sufficient *E. faecium* colonies. Samples where no *E. faecium* was isolated after sampling 20 random *Enterococcus* colonies, or where only one colony was isolated after sampling 50 random *Enterococcus* colonies, were excluded from further analysis. For the remaining index samples, colonies of *Enterococcus* were randomly sampled until 20 *E. faecium* clones per sample were isolated, requiring in some patients, isolation of up to 80 *Enterococcus* sp. colonies in order to obtain 20 that were *E. faecium*. The majority of patient samples contained multiple enterococcal species (14/22), or a combination of vancomycin resistant and vancomycin sensitive *E. faecium* (9/22). In five patients we identified only VR *E. faecium* (see S1 Appendix). For one patient (Patient 54) in the daptomycin group, only eight colonies where isolated due to low *Enterococcus* densities in the patient sample.

### Exposure and Resistance

Daptomycin exposure in the 6 months prior to the index sample ranged from 3-34 doses (days of therapy), and the most recent dose before the index sample was between 0 and 141 days earlier (see S2 Table). Patients with prior daptomycin treatment had greater proportions of resistant clones, defined as a calculated minimal inhibitory concentration (MIC_c_) greater than 4μg/ml (Fig 2). Overall 50 out of 426 clones were resistant to daptomycin with 94% of these in patients with prior daptomycin exposure. Eight of the 11 patients with resistant clones were from the daptomycin exposed group. Moreover, highly resistant *E. faecium* clones (MIC_C_ > 8μg/ml) were only found in the daptomycin exposed group. Patient mean MIC_C_ was not associated with enterococcal species diversity (Spearman ρ = -0.06, p = 0.79; S1 Appendix), the number of days since the last dose of daptomycin (Spearman ρ = -0.07, p = 0.83; S2 App), or the number of doses of daptomycin in the previous 6 months (Spearman ρ = 0.22, p = 0.50; S2 App).

**Fig 2.**
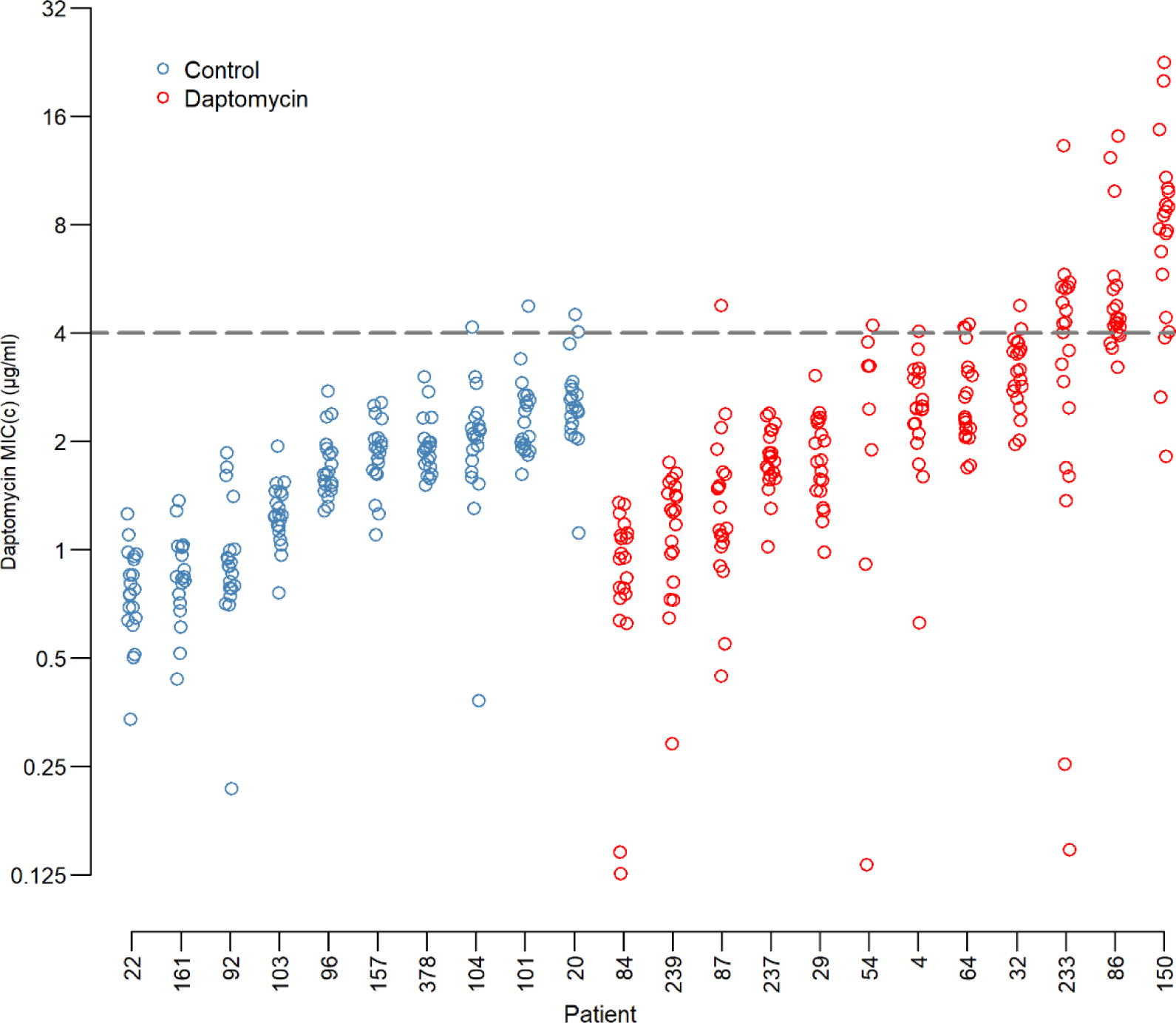
Daptomycin MICs by patient. Daptomycin resistance of each clone by patient and exposure group. Blue circles are the control patients and red circles are from daptomycin exposed patients. Each circle is the mean of two independent estimates of the MIC_C_ for a single clone. Clones above the dashed line are daptomycin resistant (clones with a calculated MIC (MIC_C_) >4μg/ml which is equivalent to the clinical 2-fold MIC cutoff of ≥8μg/ml (see Methods for a more detailed discussion of the MIC_C_ method)).

We are also interested in how variation in the resistance phenotype is spread across the study population. A Bayesian mixed-effect model was designed to test for the presence of variation in daptomycin resistance at the levels of interest: within patients, between patients, and between groups (daptomycin exposed vs not daptomycin exposed). Using the DIC criteria [24], the best fit model included a random effect for “patient”, and for “clone” nested within patient, such that the distribution for the clone effect differed from one patient to another (Table 1).

**Table 1:**
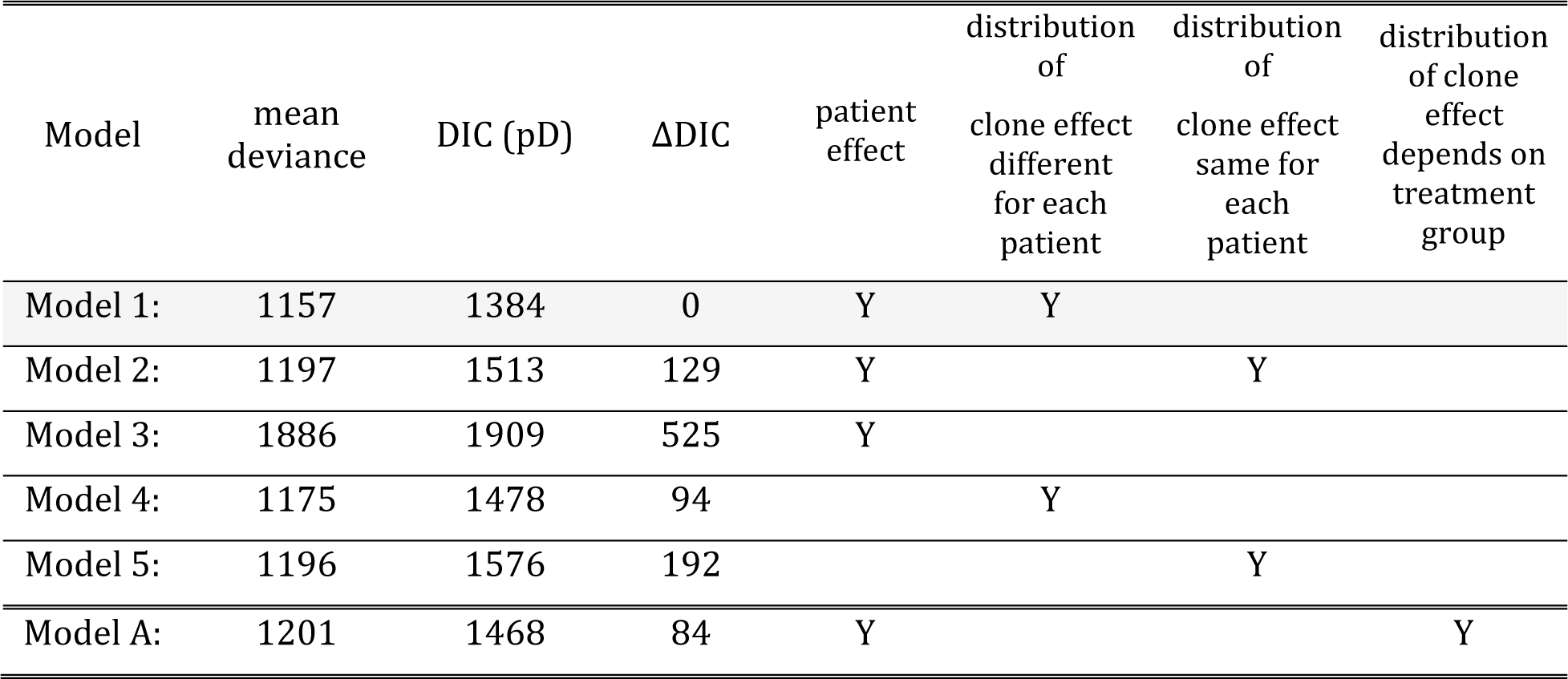
Summary of Models and DIC analysis results. Models 1 through 5 were compared using DIC criteria to ascertain how variation in resistance is spread across the study population. Models varied in the presence or absence of a patient effect and distribution of the clone effect. Model A was designed to test if clonal variation is affected by prior daptomycin exposure.

The fixed effect for daptomycin exposure, *M*_*D*,_ provided support for the hypothesis that resistance is higher in daptomycin-treated patients with 94.5% of the posterior distribution for the fixed effect falling above zero (Fig 3 insert). The mean of M_D_ is 0.57 on a log_2_ scale, which equates to approximately a 50% increase in mean MIC_C_.

**Fig 3.**
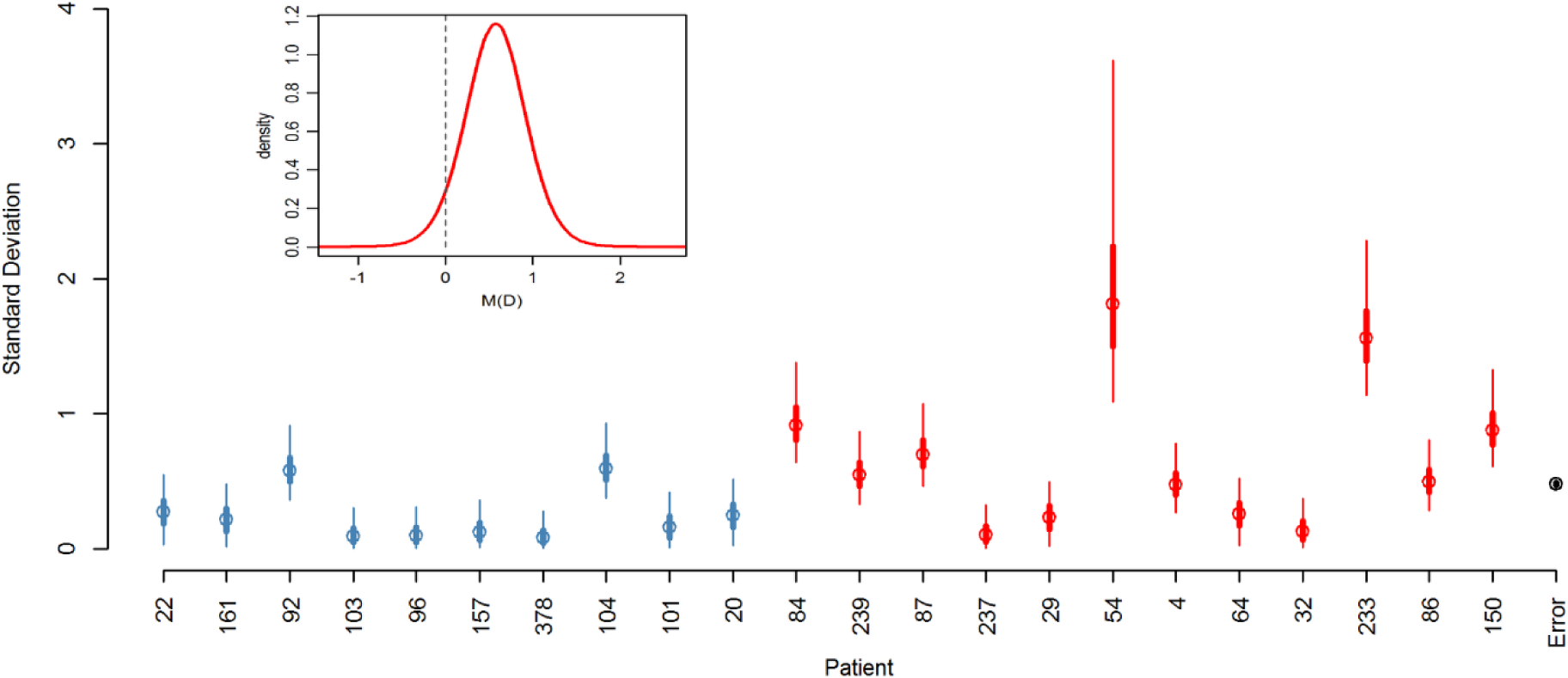
Posterior distribution of random and fixed effects from the Bayesian mixed effects model. (Main Panel) Posterior distributions for standard deviations of the clones within patients and the error. Blue markers indicate control patients, red indicate daptomycin treated patients and black is the error (Circle: mean; thick lines: 50% credibility interval; thin lines: 95% credibility interval). The 95% credibility interval of the error falls within the error marker. (Insert) Posterior distribution of fixed effect for prior daptomycin exposure.

### Within-patient Variation

The variation in MIC_C_ among clones within patients differed from one patient to the another (Table 1; Model 1 is a significantly better fit (as measured by a lower DIC) than Model 2). The posterior distributions for the standard deviations of the clone effects for each patient are summarized in Fig 3. Further, Model A was designed to assess if there was evidence that patients with prior daptomycin exposure had greater between-clone variability in MIC_C_. In particular, in Model A the standard deviation for the “clone” effect for control patients was modeled with a single parameter λ and for the daptomycin patients was λexp(*α*) (see S4 Appendix for details). By using this parameterization we were able to quantify the amount of evidence for daptomycin-treated patients having greater within-patient variability (by determining the proportion of *α*’s posterior distribution that lies above zero). There is strong evidence for daptomycin treated patients having greater within-patient variation than control patients with the estimated posterior distribution for *α* lying essentially above zero (mean=1.04, 99% credibility interval is (0.677-1.798).

### Time Series

Eight daptomycin patients and six control patients had more than one swab sample collected in 2016. While these additional swab samples do not allow for a comprehensive analysis of within-patient changes in resistance over time, they make possible a number of interesting case studies of within-patient changes in resistance. Ten clones from each of these additional samples were isolated and tested for resistance. For standardization, the first 10 clones from the index samples in the previous analysis were used.

In all three patients where a sample was collected before the commencement of daptomycin treatment, the post-treatment sample contains clones that are more resistant than any clones sampled prior to treatment. The MIC_C_ for the most resistant clone in the sample immediately prior to and post treatment are: Patient 4 prior = 2.2μg/ml, post = 4.2μg/ml; Patient 87 prior = 1.7μg/ml, post = 2.7μg/ml; and Patient 150 prior = 0.5μg/ml, post = 20.0μg/ml; Fig 4. Patients 4 and 150 also showed an increase in mean MIC_C_ (Patient 4: Prior sample mean = 1.69 μg/ml, Index sample mean = 2.86μg/ml, t = -3.46, df = 18, p=0.003; Patient 150: Prior sample mean = 0.32μg/ml, Index sample mean = 13.08μg/ml, t = -24.99, df = 18, p<0.001). Patient 87 had a higher mean in the index sample but this was not statistically different to the pre-treatment sample mean (Prior sample mean = 1.15μg/ml, Index sample mean = 1.42μg/ml, t = -1.41, df = 18, p=0.18).

**Fig 4.**
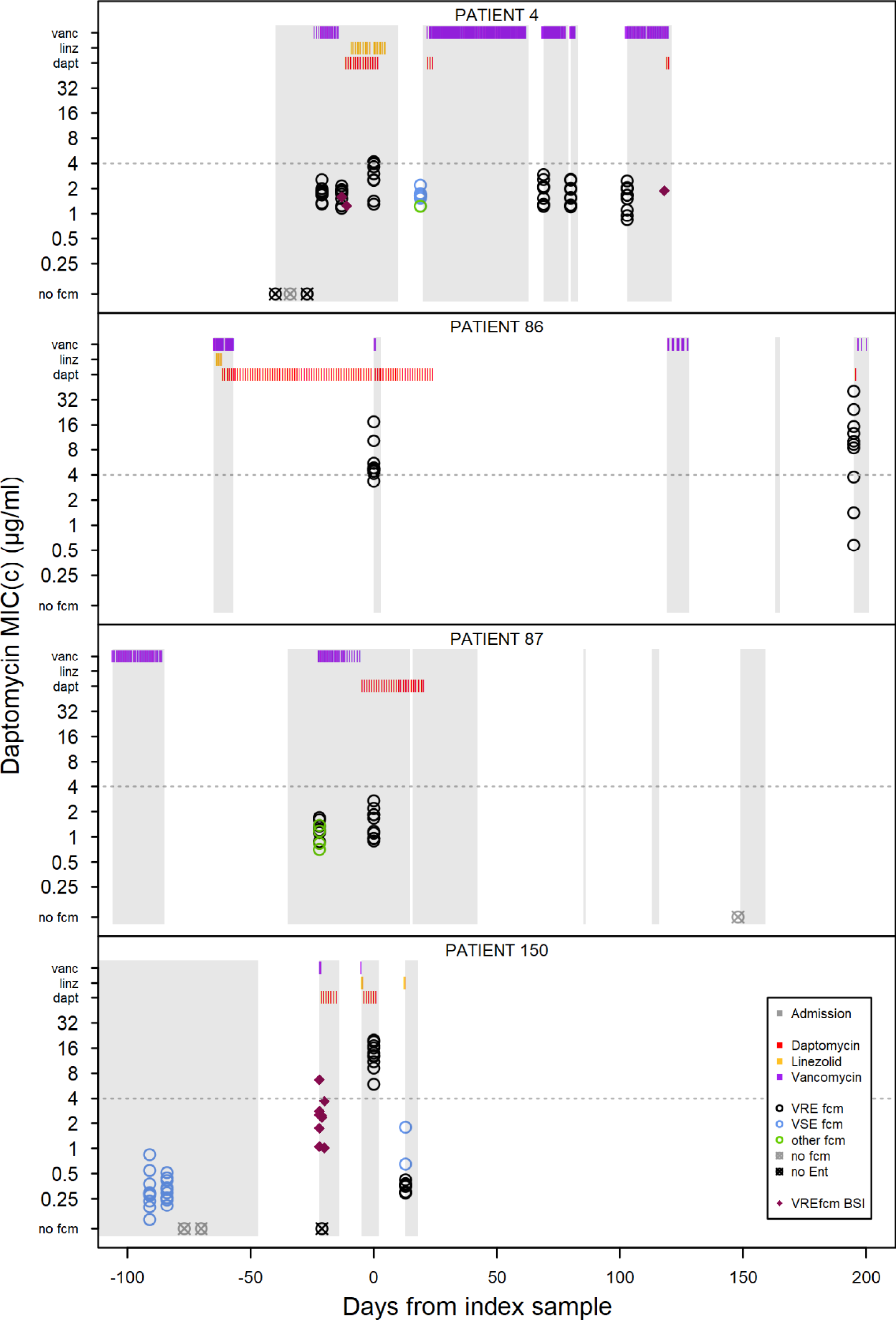
Within-patient resistance over time. Each plot shows patient admission periods, drug doses and resistance of E. faecium clones isolated from screening swabs and blood stream infections for an individual patient. Patient admission periods are shown as grey blocks, and individual doses of vancomycin (orange), linezolid (yellow) and daptomycin (red) are detailed in the bars at the top of the plot. Circles show daptomycin resistance (MIC_C_) for 10 clones per sample. Each circle is the mean of two replicates with black circles denoting VR E. faecium, blue circles denoting VS E. faecium and green circles are E. faecium with discordant vancomycin resistance genotype and phenotype. Pink diamonds are isolates from VRE faecium blood stream infections.

The maintenance of resistance following cessation of daptomycin treatment is highly variable between patients. In patients where a second sample was collected after the end of treatment (8 out of 12 patients) we see examples of long-term maintenance of resistance in the absence of further daptomycin exposure in our hospital (Patient 86) and rapid loss of resistance (Patients 4 and 150), which in both examples here is coupled with the presence of vancomycin sensitive *E. faecium* which was not sampled prior to daptomycin treatment. In two patients there was a slight increase in daptomycin resistance after the cessation of treatment which was combined with a switch from predominantly vancomycin sensitivity to predominantly vancomycin resistance (Patients 64 and 233, see S3 Appendix). In patients where the index sample contained no daptomycin resistant isolates, little change was observed after treatment ended (Patients 29 and 84, see S3 Appendix).

## Discussion

Patients exposed to daptomycin have more daptomycin resistant *E. faecium* in their intestines than unexposed patients. This difference is shown by the cross-sectional analysis (Fig 2 and 3 insert) and the patients with samples before and after daptomycin exposure (Fig 4). The findings support our hypothesis that intravenous daptomycin treatment leads to off-target selection for daptomycin resistance in the intestinal tract. Further, this finding is consistent with the ecology of this organism, with *E. faecium* most commonly found in the gastrointestinal tract and specifically adapted for transmission between patients in health care settings [25,26]. Transmission is by the fecal-oral route, which makes drug exposure and selection in the intestinal tract relevant to the risk of transmission of drug resistant pathogens [27]. Daptomycin concentrations in the gastrointestinal tract are likely in the range that would select for resistance (see below), though this exposure is evidently not enough to eradicate the bacteria. When the results of this study are added to the ecology of *E. faecium*, it is plausible that off-target selection is a major mode of daptomycin resistance evolution in hospital.

Our study also sheds light on how *E. faecium* evolves in patients exposed to daptomycin. In the absence of daptomycin exposure, few patients had detectable variation in daptomycin resistance (Fig 3). With as little as 3 doses or 3 days of therapy, the variation both within and across patients increased (Fig 3). Thus, there is either low level pre-existing variation in resistance that daptomycin exposure brings to the fore, or the supply of mutations is high enough during treatment to result in a response to selection. In either case, the response to selection is not limited by supply of resistant variants. Therefore, strategies such as mutation-prevention dosing which have the explicit goal of preventing the emergence of resistant variants may not work in the intestinal tract [28–32]. Alternative strategies that leverage competition to limit the spread of resistance may offer a way forward but often utilize lower doses [33–35], which raises the potential for conflict between controlling resistance in the gut, and controlling resistance at the site of infection.

The variation in resistance within patients was greater after daptomycin exposure (Fig 3). In fact, all patients with resistant isolates continued to also contain clones below the resistance threshold. Thus, the raw material required for the population to return to a more susceptible state remained in the gut. Determining whether this variation in resistance is associated with variation in competitive ability in the intestinal tract, or transmissibility, may help identify better resistance management strategies. However, managing resistance in the intestinal population is fundamentally different from managing it in the target population. Orders of magnitude more bacteria may be present in the intestines [36], leading to a larger pool of variation as well as increased competition, both within species, and with other members of the microbiome.

Few data are available on the pharmacokinetics of daptomycin in the intestinal tract. In healthy adult men about 5% of daptomycin is excreted in the stool over 72 hours [37]. Assuming stool water content of 100-200 ml/day [38] and 500mg daily does (6mg/kg in an average US adult) [39], the daptomycin concentration in the stool could be 125-250ug/ml, well above the MIC for most *Enterococcus*. It is unknown whether these calculations represent the drug level experienced by *Enterococcus* in the patient population in which VRE are often found, in whom liver and kidney dysfunction are common [40,41].

Temporal dynamics within patients were highly variable, even among the rather few cases with repeat sampling. In the three patients where we had a pre-treatment comparison, resistance increased following treatment. Resistance was maintained in some patients even in the absence of continued drug treatment (patient 86, Fig 4), while in other patients, populations reverted to being entirely sensitive (Patient 4: 17 days and Patient 150: 12 days; Fig 4). Thus, there are likely important unmeasured factors that influence the resistance dynamics in these patients. Likewise, it would be very interesting to know why daptomycin resistant clones were not detected in all daptomycin-exposed patients (Fig 2).

Daptomycin resistance in *E. faecium* is a growing problem that has persisted despite efforts to minimize unnecessary drug use and prevent hospital transmission [23]. This study demonstrates that off-target evolutionary dynamics likely play an important role in this problem. In systems like this, where target and non-target populations are compartmentalized, it will be challenging to identify dosing strategies that can optimally slow resistance emergence in both sites. Novel interventions that can separate treatment of infection sites from selection in off-target sites are more promising. These include interventions which neutralize the action of the drug in the gastrointestinal tract [6]. Given that many of the most serious antimicrobial resistance problems are caused by opportunistic pathogens, intervening in the evolutionary dynamics driven by off-target antimicrobial exposure in highly transmissible gastrointestinal carriage populations could have an outsized impact on the emergence and evolution of hospital-acquired resistant infections.

## Materials and Methods

### Study Participants

A case-control study was conducted utilizing perirectal swabs from an infection prevention screening program at Michigan Medicine to determine the impact of intravenous daptomycin treatment on daptomycin resistance in gut populations of *Enterococcus faecium*. The study was approved by the University of Michigan Institutional Review Board. The initial inclusion criteria (see Fig 1) were all patients with a VRE positive (*E. faecium* or *E. faecalis*) swab using VRE Select agar (BioRad) in 2016 (n=618). Patients included in the daptomycin exposure group had at least three administered doses of daptomycin (3 days of therapy) in the 6 months prior to the VRE positive swab (14 patients). The control group consisted of patients who had received no daptomycin, and at least six doses of linezolid (3 days of therapy) in the last 6 months prior to a VRE positive swab (15 patients). For each patient the first sample to meet the inclusion criteria was defined as the index sample. For each index sample, *Enterococcus* sp. clones were isolated until there were 20 E. faecium clones per sample. Samples where no *E. faecium* was isolated after sampling 20 random *Enterococcus* colonies; only one colony was isolated after sampling 50 random *Enterococcus* colonies; or where no *Enterococcus* was isolated; were excluded from further analysis. The final dataset included 12 patients in the daptomycin exposure group and 10 patients in the control group. For a further time-series analysis of these patients up to 5 prior samples and all subsequent samples available from these patients in 2016 were tested for the presence of *E. faecium*. 10 random clones per sample were isolated from each of the *E. faecium* positive samples. Finally, we collected isolates from all blood stream infection (BSI) in these patients.

### Ethics Statement

This study was approved by the University of Michigan Institutional Review Board (ID no. HUM00102282), which determined that informed consent was not required as all samples utilized were collected for patient treatment purposes.

### Strain Isolation

Perirectal swabs were obtained using E-swabs (BD) as part of the hospital VRE surveillance program. The swabs were first tested in clinical microbiology lab by streaking on VRESelect agar (BioRad) per manufacturer’s recommendations. The swab was discarded and the residual media was stored with glycerol (final concentration 20% v/v) at -80°C. To isolate *E. faecium*, samples were streaked from the sample stored in glycerol onto Enterococcosel agar (BD BBL) in duplicate and incubated up to 72hrs at 37°C. The first 10 colonies from each plate (20 colonies per sample) were re-streaked on Enterococcosel agar and incubated for 48 – 72hrs at 37°C. One colony from each plate was then streaked on BHI agar (BD BBL) and a vancomycin (30μg/ml Oxoid) disc was placed on each plate to determine vancomycin resistance. Plates were incubated for 24hrs at 37°C. One clone per plate was stored in BHI +20% glycerol at -80°C.

To confirm the species of *Enterococcus*, a species-specific multiplex PCR was performed using primers for *E. faecium, E. faecalis* and VanA, VanB, VanC1 and Van C2/3 [42]. Briefly, 11.25µl PCR Master Mix (iProof HF, BioRad), 50uM of each primer and 6.45µl water (total volume 22.5µl) per sample were combined. Sample was added as either 2.5µl of bacteria in BHI glycerol taken from tubes prior to freezing, or 1 colony from a streaked culture of stored bacteria. PCR was performed under the following conditions: 95°C for 4 mins; 30 cycles (98°C for 10 secs, 55°C for 30 secs, 72°C for 30 secs); 72°C for 7 mins. Gels were run for 1hr at 80-100V on 2% Agarose in TAE buffer with 0.1µl/ml SybrSafe.

Isolation steps were repeated until 20 *E. faecium* clones per sample or 10 clones for time-series only samples were isolated. Samples were excluded if more than 40 *Enterococcus* clones were isolated without any *E. faecium* (20 clones for time-series only samples), or no *Enterococcus* was detected after streaking the sample twice and then plating 80μl of the initial patient sample on Enterococcosel agar (combined ∼10% of the total sample volume). This sampling method resulted in a dataset on species diversity within patients (see S1 Appendix).

Ten patients had *Enterococcus* blood stream infections (BSI) within six months of the index swab sample, and a total of 45 isolates were taken from these patients. Blood samples were cultured in blood bottles and streaked on Chocolate agar in the clinical microbiology lab. Single colonies were streaked on BHI agar three times and then a single colony was stored in BHI +20% glycerol.

### MIC testing

MIC testing was performed by the broth microdilution method (BMD) according to CLSI M7 guidelines [43], each samples was tested in duplicate and one of four patient-derived *E. faecium* strains was included on each run as a positive control. All clones were initially tested on plates containing 2-fold dilutions of daptomycin with final concentrations ranging from 0.125µg/ml to 16µg/ml and the optical density (OD) of each well (600nm) was determined by plate reader (FLUOstar Omega, BMG Labtech). OD values for each dilution series were fitted to a Hill function to determine the concentration at which the hill function curve crossed the cutoff (defined as 2SD above the mean of the negative control wells (see S4 Appendix), we refer to this value as the computed MIC (MIC_C_). If the initial concentration range did not contain at least two concentrations above and below the cutoff, the assay was repeated on either increased (1µg/ml to 64µg/ml) or decreased (0.0625µg/ml to 4µg/ml) concentrations as appropriate. Individual assays of clones were also excluded if the Hill curve did not fit the data points well, determined as an MIC_C_ greater than one 2-fold dilution from the lowest concentration with an OD below the cutoff.

### Statistical analysis of *MIC*_*C*_

We analyzed the log_2_ of the MIC_C_ values using Bayesian mixed effect models [44]. Models included a fixed treatment effect for patients that fulfilled the daptomycin exposure case definition (see above). To quantify the evidence for prior daptomycin exposure increasing MIC_C_, we determined the proportion of the posterior for the fixed “treatment” effect that was above zero. The full model included 23 random effects (one ‘‘patient” and 22 “clone” effects) and allowed the distribution of the “clone” effect to depend on patient. We fit six candidate models which considered different combinations of the random effects. In addition to testing different combinations of the random effects from the full model, we also considered models where the distribution of the “clone” effect was identical for all patients, and a model where this distribution depended on treatment group (see Table 1 and S4 Appendix for details). All random effects are assumed to be normally distributed with zero mean and standard deviations estimated using the MCMC program JAGS [45,46]. Uninformative priors were used (see S4 Appendix Table 2). We ran each model for 20 × 10^6 iterations with a burn in of 10×10^6 steps and a thinning interval of 2 × 10^3. This resulted in 5×10^3 parameter samples for each model run. This process was repeated to generate four chains, with randomly chosen initial starting values, for each model. Posterior convergence was confirmed in two ways: i) empirical inspection of the estimated posterior distributions (all four chains resulted in very similar distributions) and ii) the Gelman-Rubin convergence diagnostic (this statistic was essentially 1 for all parameters, which is consistent with the chains having converged).

The models were then compared using the Deviance Information Criterion (DIC) [24]. The relative fits of the models are summarized using ΔDIC scores which are the differences in DIC between the best model and each alternative model. Although there is no universally agreed threshold for significance of ΔDIC scores, there is precedent for treating ΔDIC scores greater than 10 as providing very little support for the model [47](similar to rules used for the Akaike Information Criterion (AIC) [48,49]). The smallest ΔDIC score was 84, indicating that Model 1 is clearly the preferred model. To assess how well the data fit the best model (as selected by DIC comparisons), we used the posterior distributions of the selected model to generate synthetic data sets and examined the distributions of a number of different summary statistics (see S4 Appendix).

## Data Availability

Data will be made available upon publication of the manuscript. For access to data prior to publication contact the corresponding author.

## Acknowledgements

We thank Amit Pai for discussion about pharmacokinetics.

## Supporting Information Captions

**S1 Appendix. Sample Species Diversity** Distribution of *Enterococcus* species with patient samples.

**S2 Appendix. Timing and number of doses of daptomycin prior to index sample**.

**S3 Appendix. Time-series Plots for All Patients**

**S4 Appendix. Supplementary Methods and Analysis**

